# Evaluation of three control strategies to limit mpox outbreaks in an agent based model

**DOI:** 10.1101/2024.02.06.24302176

**Authors:** Julii Brainard, Iain Lake, Paul R. Hunter

## Abstract

Most of the 2022 mpox outbreaks in high income countries, which predominantly affected men who have sex with men, peaked less than two months after detection. To stop the outbreaks, people were encouraged to limit new sex partners, take up any offers for smallpox vaccination, and self-isolate. The relative contributions of each of these strategies to outbreak reduction are hard to know. To consider the potential relative efficacy of each of these measures individually, we constructed agent-based models using plausible partnership counts, reasonable behaviour choices and published information about smallpox vaccination uptake rates in the UK context during 2022. Compared to a baseline, no intervention scenario, partner reduction was more effective at preventing generation of secondary cases than the vaccine rollout at the speed that the smallpox vaccine rollout occurred in the UK in 2022. These findings suggest that partner reduction by the most affected community rather than pharmaceutical intervention was largely to credit for causing case numbers to peak as early as they did.

## Introduction

Mpox is a smallpox-like zoonotic infection caused by a virus in the Orthopoxvirus genus originally endemic in some areas of Africa. The most common mpox symptoms are fever and lymphadenopathy with many persistent and potentially very painful lesions. Recovery typically takes 2-4 weeks and confers long-lasting immunity to reinfection. Transmission to humans occurs after physical contact with an infectious individual, contaminated materials or an animal ^1^. The historic case fatality rate was thought to be about 10%, and was known to be higher in children and lowest in healthy adults ^2^. Although no vaccine has been developed against mpox specifically, smallpox vaccines have high efficacy against mpox, reducing disease acquisition by about 80% after one or two doses ^3,4^. It is thought that protection from the smallpox vaccination may decline over time, but only slowly ^5^. That cessation of routine community vaccination against smallpox might lead to recurring mpox outbreaks was suggested ^5^.

Historically, most mpox outbreaks were quickly contained and seemed to involve relatively short transmission chains (under 7 generations before outbreak end ^2^. However, a large number of mpox cases in new countries and continents were identified in 2022, causing fears that the disease might become endemic in many new regions. The 2022 multi-country outbreak overwhelmingly affected men who have sex with men (MSM)s ^6^. Most hospitalisations in high income countries in 2022 were due to need for analgesic relief ^6,7^, and relatively few deaths were associated with the 2022 multi-country outbreaks ^8^ A modest observed case fatality rate (CFR, probably < 1%) in 2022 is thought to relate in part to the specific (milder) clade 3 viruses that the 2022 outbreak cases mostly have^9^ as well as under-ascertainment of cases where mpox was traditionally endemic which may have skewed historical estimates of the CFR. However, to prevent onward disease transmission, even non-hospitalised cases were strongly advised if not also legally required to strictly self-isolate for a minimum of three weeks, causing economic and social disruption for those affected.

Most of the 2022 mpox outbreaks in high income countries peaked less than two months after detection. To bring the outbreaks under control, MSMs were encouraged to limit new sex partners, to take the smallpox vaccination, and to self-isolate if they knew they had the virus. The relative contributions of each of these strategies to outbreak reduction are hard to know. Contact tracing was expected to be challenging because of privacy worries, relatively high incidence of anonymous pairings and social stigma ^10^, making it difficult to find people who needed to be warned to be vigilant and seek testing if they had relevant symptoms. In part because of limited smallpox vaccine supplies, vaccination programmes in most settings were not offered universally and instead initially targeted MSMs thought to be at greatest risk of infection due to behavioural history. Partner reduction and vaccination programmes had apparent success, in that by mid August 2022, about 50% of American MSMs reported reductions in their sexual activity to prevent catching mpox ^11^, and by mid-November 2022 take up of the smallpox vaccine among eligible UK-resident MSMs was near 50% ^4^. The level of adherence to self-isolation guidance has been harder to ascertain. Self-isolation for persons with confirmed infection or suspected exposure was encouraged (for instance, in Britain from 22 May 2022 ^12^) but not legally required in many jurisdictions.

Nevertheless, it seems likely that behaviour changes (partner reduction and compliance with guidance to self-isolate) as well as vaccination programmes all contributed to outbreak reduction. To consider the potential relative efficacy of each of these measures in isolation, we constructed agent-based models ^13^ for mpox transmission among MSMs using plausible partnership counts, reasonable behaviour choices and vaccination uptake rates. The findings were used to suggest which of three common mpox control strategies may have made the greatest contribution to early subsidence of the outbreak. Sensitivity analysis was undertaken by varying the most uncertain epidemiological parameters.

## Methods

### Overview

Baseline and alternative scenarios were constructed within agent based models (ABMs) using Netlogo version 6.0.2. Agents in the models had four possible states that occurred in this sequence: susceptible–exposed−infected−recovered (recovered = immune; SEIR). All of our simulations consider the case of 6400 agents who are all MSMs, with no new members added or departing during each simulation (a closed system). We used a population of 6400 agents in order to be able to generate many simulation results in a reasonable time frame (larger agent counts would have taken much longer to achieve modelling results). Outcomes are always monitored from simulation start (when control measures start to be applied) until outbreak end (when no more agents are infectious or incubating).

We identify plausible parameter estimates and assumptions, specifying our preferred estimates and assumptions, treating the adoption of these plausible assumptions as a baseline and reasonably plausible scenario. We then explore what happens if we use alternative credible estimates or assumptions in other key epidemiologic parameters. First, we consider the posited control strategies under the baseline conditions. We also varied epidemiological and control parameters, to assess sensitivity to selected assumptions.

We compare outbreak control with respect to two harmful outcomes: duration and number of new cases. Outbreak duration is defined as duration in hours or days from simulation start (when control strategy starts to be applied) until there are no more infectious or incubating agents (outbreak end). Number of new cases is defined as the number of new cases that occur after the simulation start until outbreak end. We report outcomes after 1000 simulations for the baseline scenario and suggested plausible implementations of each control strategy. As sensitivity analysis, we run 100 iterations for alternative ways of implementing each control strategy with varying assumptions about transmission risk per contact different numbers of seed agents. The outcomes do not account for hospitalisations or deaths because we were interested in scenarios when most symptoms were mild, mortality rates very low and most cases stayed in the community, similar to most of the 2022 outbreaks. We do not consider the possibility of asymptomatic transmission because it seemed unlikely; a 2022 audit of 2917 specimens from MSMs attending sexual health clinics in England found low prevalence (0.14%, n=4) of non-symptomatic mpox infection ^14^. We do not consider the possibility of airborne transmission because of previous models which suggested that this was likely to be relatively rare for mpox ^15^.

### Fully fixed model parameters

ABMs incorporate a blend of stochastic and deterministic parameters. There are fixed (deterministic) aspects of our models (same in all scenarios), partly deterministic and partly stochastic parameters, as well as explicitly varied parameters (for sensitivity analysis).

We fully fixed aspects of our modelling environment for pragmatic reasons and/or where the effect of varying the assumption was predictable. Fully fixed aspects of our models in all simulations are: count of agents (6400), population under study are all MSMs and the system is closed : there are no new members, no agent departures and no new sporadic case introductions. The choice of 6400 agents is not meant to be definitive or a priori known to be optimal. Rather, we use a fixed number of agents in the models to standardise the scenarios relative to the sensitivity analyses, such that the factors that seem to cause changes in the primary outcomes (duration of the outbreak and new case counts) should arise from variations fundamental to the control strategies tried rather than variations in the population size. It is predictable that a larger population would have potential for longer duration outbreak, so we do not undertake sensitivity analysis by varying the total number of agents. 6400 agents for the scenarios is also a pragmatic choice to enable extensive sensitivity analysis of other factors, and many simulations and models to be generated and summarised using our available resources within a feasible timeframe.

Similarly, the difference between closed and open populations on outbreak development is predictable, an open system (new agents enter and some agents depart) is likely to have longer duration and more variable development. An outbreak or epidemic in a closed system will always come to an end, as long as reinfection is not possible, assuming that new sporadic cases cannot occur. We also confined our modelling to account for just one kind of sexual partnership (men who have sex with men, MSMs); the 2022 outbreaks in novel settings overwhelmingly stayed within this sexual minority community. Adding other sexual preference groups (eg., women who have sex with men or women who have sex with men who have sex with men) would have added complexity to our own models without increasing insight into which mpox control strategies had greatest potential to work well in a predominantly MSM outbreak.

### Model assumptions that were mixed deterministic and stochastic

Aspects of the agent interaction environment had fixed attributes (such as median value for entire population) but these values varied for individual agents between scenarios, thus these environmental and behavioural model assumptions are partly deterministic and partly stochastic.

#### Homophily in sexual contact network

A key aspect of model construction, to make sure the model represented realistic behaviour choices, was to establish homophily within social networks. Homophily is the principle that people tend to socialise with other people like themselves ^16,17^, and in this case for sexual partnership networks, with other persons who have similar sexual appetites to oneself. To achieve homophily, one third of agents were located at random places in the agent world. Most (target 67%) agents were located (stochastically in individual model runs) in 64 clusters with similar sexual appetites, initially placed on model development to follow proximity and density rules used in authors’ previous ABMs ^18^ to achieve contact rates and relationship contact similar to those reported for the UK in observational research ^17,19^. A gamma distribution was used for expected sex act distribution, corresponding with the ‘heavy tail’ observed in some MSM sexual contact networks ^15^.

How strong homophily (on sexual appetite dimension) is within the MSM community was not feasible for us to parameterise with empirical data, rather we created variety in homophily (similarity for individual agents with others in their contact circles) that was plausible. Clusters haddimensions that were approximately 12.2 distance units diameter. The median number of agents within a 6.1 radius of each agent was about 144 in most model runs (range approximately 42-242). Within this 6.1 radius circle, about 67% of agents had an average of half of their contacts with similar sexual appetites; remaining agents within 6.1 distance units had a diversity (stochastically placed) of sexual appetites. In this way, some agents ended up with high homophily in their local area; others had high variation in sexual appetite / low homophily among their social contacts. Ultimately, we assessed the performance of our models and agents’ behaviour within our models, against prespecified and plausible targets about how many unique partners and unique sex acts that agents typically experienced in model runs (see Table 1).

**Table 1.** Scenario Descriptions.

| *Control strategy* | *Scenario description* | *Details* |
| --- | --- | --- |
| Casual partnership reduction | High | 50% of agents cease casual partnerships |
|  | **Plausible** | **50% of agents cease 50% of casual partnerships, ie., 50% of such opportunities still happen** |
|  | Low | 50% of agents cease a small % (20%) of casual partnerships |
| Vaccination programme | Low: 25%<br>**Plausible 50%**<br>High: 75% | Speed of vaccine delivery; % of highly at-risk target group that receive vaccination by day 120 |
| % who Self-isolate after diagnosis | Low 11%<br>**Plausible 22%**<br>Higher 33%<br>Maximal 44% | % of infected agents who heed advice to stop all sexual contact when infectious (after potential delay to realise own case status) |

Note: Assumed uptake of vaccine is constant (same number of doses delivered daily) accumulating to 50% of the group offered the vaccine, by 120 days after vaccination programme starts.

#### Calibration and partner seeking behaviour

Another important objective of the cluster placement and indicators that the models were representing partnership behaviour as intended, was to ensure that most agents had low partner counts (≤ 1 over a three week monitoring period), and that most sex acts were with a limited number of other agents, resulting in the prespecified target counts of unique sex acts and unique partners (see Table 1). Density of agent placement and distances moved each hour (when they could plausibly encounter new agents), as well as search radius from which to select a potential partner, were empirically calibrated in model development, as a function of sexual appetite. Adjustment of travel and search distances allowing for sexual appetite was to ensure that the sexual activity (in terms of unique sex acts and partner counts) of agents fell on the target distribution over 3 week periods (median partner count = 1, median sex acts = 3-6), and that higher unique partner and sex act counts would result for the agents with highest sex appetite. The models had a further target that the maximum sex acts achieved should tend to have median about 32 acts/week, and unique partner count with median maximum ≤ 24. These targets guiding model development and calibration are meant to be plausible not definitive. The target for median (1) unique partner counts was chosen with reference to empirical UK data collected about partner counts over three week periods, which data oversampled from MSMs using digital applications designed to facilitate casual sexual encounters, in August-September 2022 ^20^.

For comparison, 53% of 5193 American MSMs in 2009 reported that they had sex with a male partner less often than once every three weeks, while 14.1% reported having sex with another male at least twice a week. We thought it was preferable that agents in our models were calibrated to be more sexually active than the general population of MSMs, because we wanted to include more high risk individuals, and thus our agents had targets of median 3-6 sexual acts per 3 week monitoring period.

All agents return to a ‘home’ location at end of each 16 hour waking period, this leads to higher contact rates with the same agents. A majority of agents (target 60-70%) have a single preferred partner who is in close proximity to home-location (within 1.48 distance units of home location), the “steady”, and thus should be encountered often. The target (60-70%) corresponded with an observed 66% (106/161) MSMs surveyed from the general population who reported having a partner in a publicly available dataset of gay, bisexual and other MSMs surveyed in September 2022^21^.

Each waking hour (16x/day) each infectious agent is queried for their willingness to have sex, which willingness is a function of their fixed sexual appetite attribute. Empirically, the models are designed such that the median number of sex acts in a three week period should be 4-6 and the median number of unique partners (during the infectious period, which also has median duration of about three weeks ^15^) should usually be 1. Most agents strongly prefer to have sex with their steady if they have one, rather than other partners who might be available. The steady does not change during the period when an agent is infectious. The steady relationship is 100% reciprocal, but the inclination to have more than one concurrent partner, to seek partners in addition to the steady (“stray.threshold”) is not reciprocal. The population median stray.threshold was set to about 11%. That threshold % was determined empirically to create several target model outcomes: that > 50% of all sexual acts for all persons who have a steady, should be with the steady; that maximum partner count (for any individual agent) during infectious period should ≤ 24 and maximum sex acts during infectious period be ≤ 32.

#### Duration of period when infectious

Most sources agree that the infectious period (which ends when scabs have fully dried and dropped off) tends to have 2-4 week duration after symptom onset ^15,22^. USA CDC guidance is that scabs form on days 7-14 days of mpox illness and tend to take about a week to crust over and start dropping off, but infectiousness is not considered likely to have ended until skin has completely healed over after the scab fell off ^23^. Mpox patients have tested positive on PCR as late as day 39 of illness ^24^. However, PCR confirms presence of at least some viral DNA fragments rather than confirms that the DNA is activated (can replicate and is therefore infectious); testing positive on PCR after the infectious period has ended is common ^25^. With these data in mind, similar to other models ^15^, we assumed that the infectious period had median duration 21 days (504 hours), with range 14-28 days. The infectious period for individual agents is stochastic within that range, with a Gaussian distribution around the median. Full immunity follows after infectious period finishes.

#### Daily contact cycles

Models always start at beginning of day (nominally 7am) with 16 active hours considered for disease transmission opportunities, this means the maximum number of sex acts per day that could happen is 16. To confirm that models were constructed as specified, we monitored several parameters such as how many unique partners and unique sex acts were typically achieved by infectious agents during their infectious period (approximately three weeks).

### Epidemiologic assumptions varied in sensitivity analysis

We undertook sensitivity analysis varying some model assumptions and each of the control strategy options. In our baseline plausible model at simulation start, the number of infected agents (able to transmit disease to others) numbered 16, which could correspond to a detected case count of 8 and undetected cases numbering 8. As alternative start points for outbreak control, we considered the case of 8, 32 or 64 initially infected agents in sensitivity analysis. Remaining agents were assumed to be susceptible with no relevant vaccination history.

The real risk of mpox transmission per sex act (or per type of sex act) is unknown. Early (data collected 1981-1986) estimates of secondary attack rates (SAR) for mpox were as low as 3% ^26^. However, mpox SAR estimated since 2015 for persons in same households with index cases (not necessarily between sexual partners) have been much higher, 50-60% ^27,28^. In sensitivity analysis to see if findings about control strategies were dependent on the transmission risk assumptions, we consider six fixed transmission risks (per sexual encounter): 6%, 12%, 24%, 36%, 48% and 60%. In our preferred baseline model we apply the expectation that there was a 24% risk of transmission each sex act from infectious to susceptible. Our models don’t allow for variation in transmission risk that may depend on specific sexual acts.

### Control Strategies

Three fundamental control strategies are compared. Each control strategy was considered individually (not in combination with other control strategies). The control strategies were described and applied with variations for sensitivity analysis in assessing their inherit potential. It was obvious that combined strategies would be more effective than any strategy in isolation so we did not generate models to describe the obvious advantages of deploying multiple control strategies in combination. Rather, our interest was in considering plausible individual potential of each control strategy. Table 1 summarises the scenario descriptions to accompany the descriptions below.

#### Scenario 1: Partner reduction

In this scenario, there is a reduction in the proportion of sex acts that are casual (casual means no steady partner to consider) or concurrent (strays from a steady partner). About 50% of American MSMs surveyed in 2022 reported behaviour to reduce their sex partner count in order to prevent catching mpox ^11^. It was unclear how to reflect this reported behaviour change in agent interactions. We opted for three different interpretations, enabling sensitivity analysis in the evaluations of a partner reduction strategy. A maximal interpretation is that 50% of all agents stopped all casual and/or straying sex during the outbreak duration, which is very much like our self-isolation scenarios (below) so we don’t model this maximal interpretation as part of Scenario 1. Another possible interpretation might be if 50% of agents consistently reduced their casual/straying partnership count by 50%, and the other 50% behaved as normal. A more widespread but less consistent partner reduction strategy which we treat as our most plausible and preferred interpretation, could be if agents randomly reduced their casual/straying partnerships by 50% (50% of such opportunities still happened). A third possibility is that 50% of agents behaved as normal, while casual/straying partnerships reduced for 50% of agents, but the reduction was small and effected by random agents (80% of such events still happened but 20% did not).

#### Scenario 2: Vaccination only

In this scenario, vaccination uptake is high and effective, but there are no behaviour changes. The model assumes that the vaccination protection is immediate but only if received before exposure: vaccinating an already infected agent does not reduce subsequent infectiousness. Otherwise, the level of protection from vaccination in the models was stable (does not decline over time). Recent data indicated that a single vaccine dose caused 78% of recipients to become immune to mpox ^4^. Vaccination in the model is delivered as a single dose, targeted at individuals who have the highest risk of catching mpox.

Deciding how many of our agents should be offered the vaccine was a tricky decision. Actual offer in Britain was probably to only about 15% of resident MSMs. We estimate 15% because there was almost 50% uptake to those offered, reported as 55,000 first doses of the smallpox vaccine that were administered to MSMs in Britain in the 4 month period leading up to 3 Nov 2022 ^11^, which compares to an estimated 754,000 British men who identified as gay or bisexual in 2019 ^29^, and an unclear number of British men who identify as heterosexual but are also MSMs. Only about 10% of agents in our baseline model finish with zero partners over a three week monitoring period. This compares to about 35% of > 2000 general population MSMs in a September 2022 survey ^30^ who reported they had zero partners in the preceding three weeks. However, real world partner counts reported by MSMs in 2022 may have been much diminished because of concurrent mpox awareness (real life scenario 1 decisions), making the real-world survey partner counts also unrepresentative of uncontrolled behavioural preferences. We came to the pragmatic decision that in our preferred baseline model, about 30% of our agents (those at highest risk) would be offered the vaccine. We varied the pace of delivery, however, such that the accumulated delivery was 25%, 50% or 75% of target population by day 120 of strategy implementation. We deemed as most plausible scenario, that 50% of the group offered the vaccine had uptake by day 120.

High risk individuals were defined as agents who are A) still susceptible and B) either/both among the agents with highest sex appetite and/or agents with highest tendency to practice partner concurrency (straying attribute; some agents were in both groups). These high risk agents received at random a single smallpox vaccination. This mean for instance, where 50% were vaccinated by day 120, that 39% of the entire high risk target group would no longer be susceptible to mpox, because of their vaccination. In practice, because the model started with exactly 6400 agents, the high risk group tended to number about 1920 and the number who should be effectively protected by vaccination if 50% delivery target achieved after four months was about 749 (∼39% of 1920): we monitored model parameters to make sure this outcome was achieved.

#### Scenario 3: Some self isolation

We have not found data describing actual adherence to self-isolation advice after mpox diagnosis in 2022 in the UK. 44% of Dutch MSMs surveyed in July 2022 ^31^ indicated high willingness to self-isolate if they had mpox, at least until all lesions were gone. Given that stated intentions to comply with public health advice tend to be much stronger than actual compliance ^32^, the percentage of persons with confirmed mpox diagnosis who have actually fully self-isolated has probably been much lower than 44%. In sensitivity analysis we tested the scenarios where either 11%, 22%, 33% or 44% of infectious agents isolated after their symptoms start. We chose 22% as the most plausible value. However, self-isolation start was assumed to often be delayed to reflect likely delayed recognition/acceptance of own case status. For those who self-isolate at all, delay to start self-isolation follows a gamma distribution (long right tail) after first signs of infection, median delay is about 72 hours, and range 0 to 400 hours. Other agents don’t self-isolate at all when infectious.

## Results

Table 2 shows model targets and values achieved, with regard to agent behaviours and outcomes. The model achieved most of the behaviours and agents had qualities consistent with the model design objectives. Table 3 shows outbreak outcomes (medians and IQRs) for the baseline and preferred alternative scenarios. Tables S1-S4 in Supplementary Material show outbreak outcomes varying epidemiologic parameters with each of the candidate control strategy assumptions.

**Table 2.**
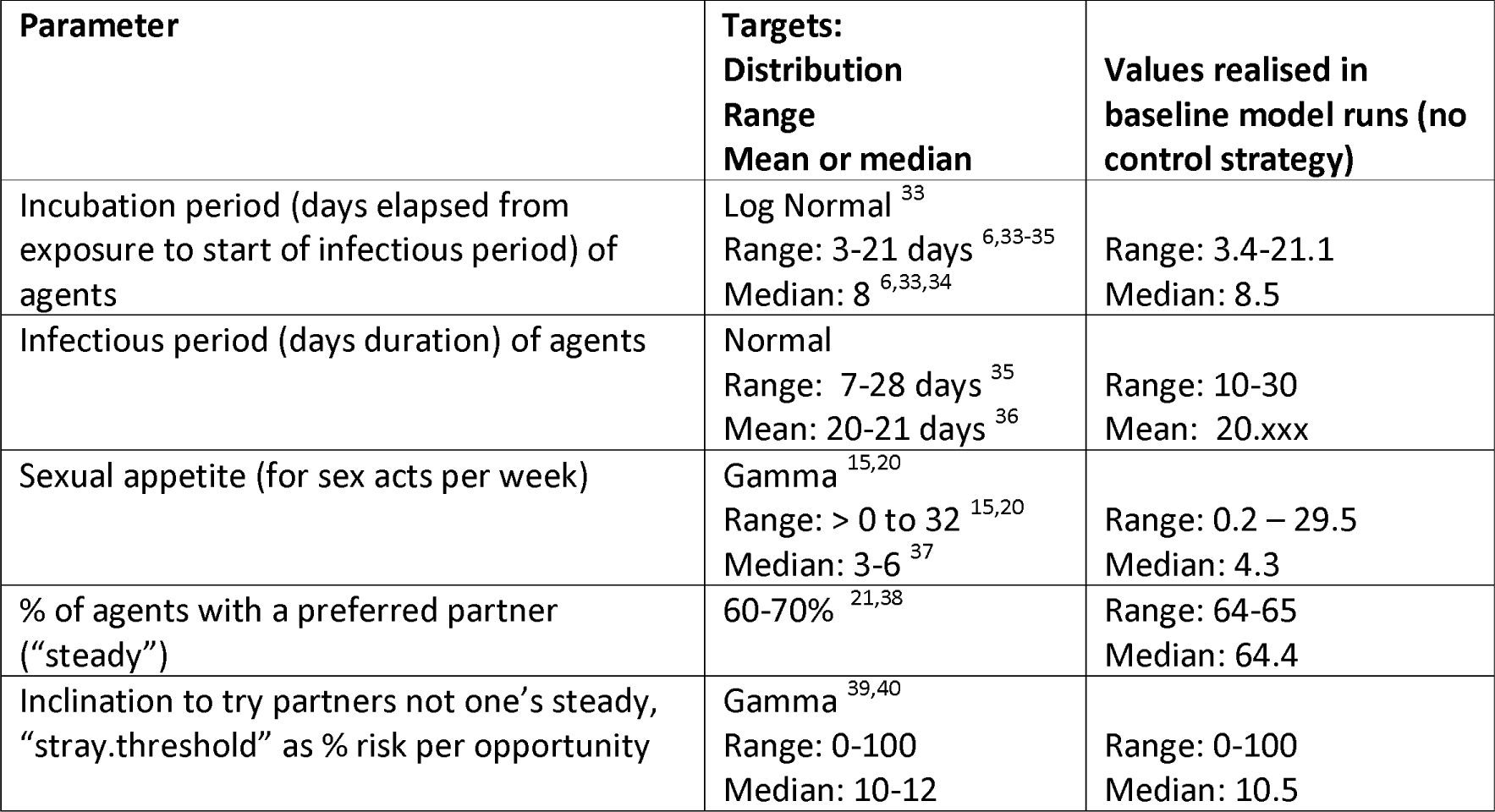
Verifying model performance in baseline scenario: stochastic assignments.

| Parameter | Targets:<br>Distribution<br>Range<br>Mean or median | Values realised in<br>baseline model runs (no<br>control strategy) |
| --- | --- | --- |
| Incubation period (days elapsed from exposure to start of infectious period) of agents | Log Normal<sup>33</sup><br>Range: 3-21 days<sup>6,33-35</sup><br>Median: 8<sup>6,33,34</sup> | Range: 3.4-21.1<br>Median: 8.5 |
| Infectious period (days duration) of agents | Normal<br>Range: 7-28 days<sup>35</sup><br>Mean: 20-21 days<sup>36</sup> | Range: 10-30<br>Mean: 20.xxx |
| Sexual appetite (for sex acts per week) | Gamma<sup>15,20</sup><br>Range: > 0 to 32<sup>15,20</sup><br>Median: 3-6<sup>37</sup> | Range: 0.2 – 29.5<br>Median: 4.3 |
| % of agents with a preferred partner (“steady”) | 60-70%<sup>21,38</sup> | Range: 64-65<br>Median: 64.4 |
| Inclination to try partners not one’s steady, “stray.threshold” as % risk per opportunity | Gamma<sup>39,40</sup><br>Range: 0-100<br>Median: 10-12 | Range: 0-100<br>Median: 10.5 |

**Table 3.**
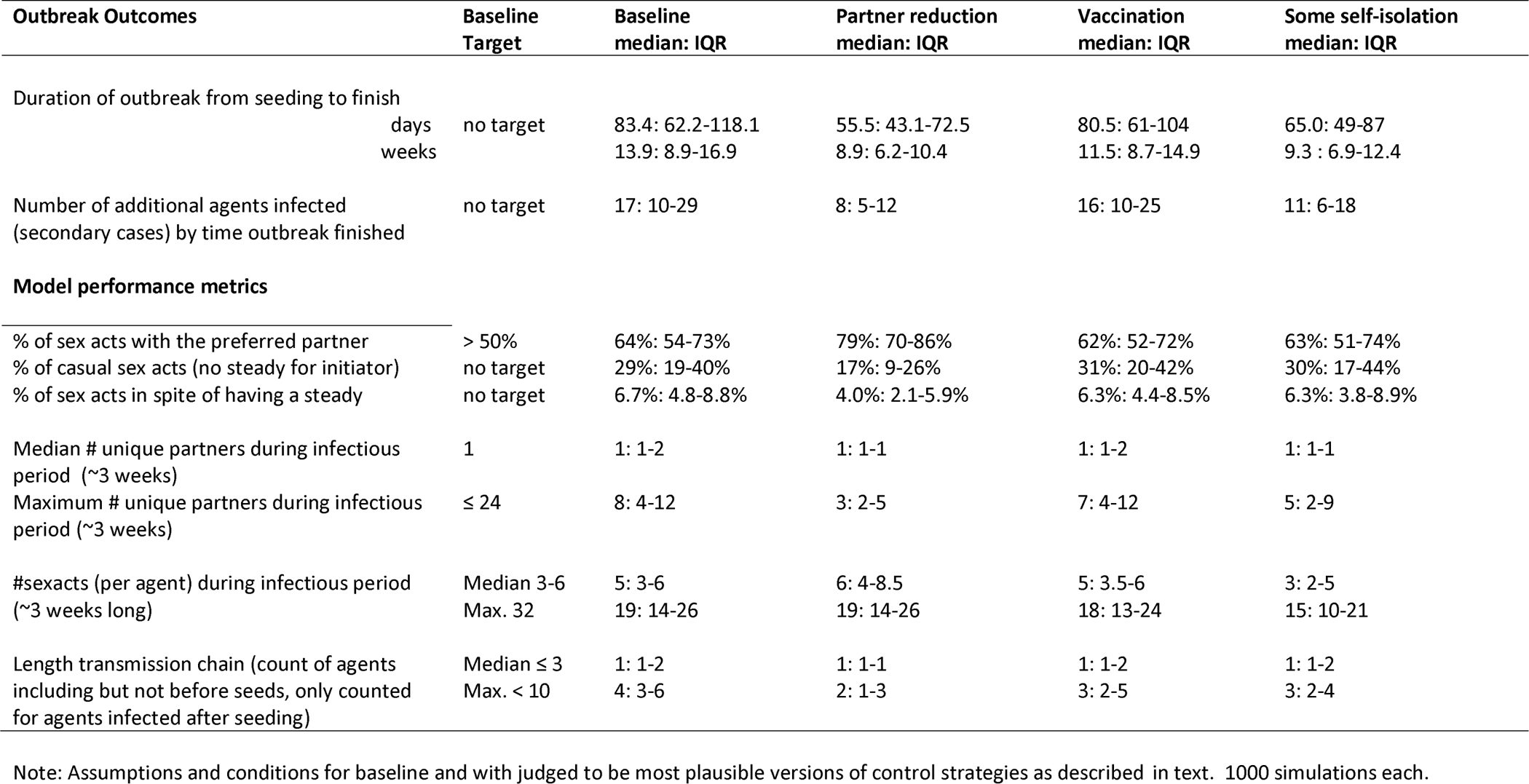
Model performance and outbreak outcomes observed in each simulation, under baseline conditions and plausible control strategies; without sensitivity analysis.

Monitoring of model behaviour, parameter assignments and some outcomes all indicate that the models performed as designed. For instance (Table 3) 67% of agents are strongly in contact clusters, so they will tend to encounter the same other agents most commonly. Similarly, incubation periods, recovery/infectious periods and other parameters are assigned to agents on the desired distributions (Table 1). The median number of partners is 1 in most simulations, while the transmission chains (counts of generations in the mpox outbreak) are < 10 consistently (Table 3).

In addition to behaviour outcomes, Tables 3 and S1-S4 report the key epidemic outcomes under preferred plausible baseline and control scenario assumptions: outbreak duration and total cases generated after simulation start for baseline and under some plausible control strategy implementations. In Table 3, reducing casual encounters partner concurrency (Scenario 3, 50% of such incidents don’t happen) leads to the most improved (lowest) count of secondary cases and results in the shortest duration, from median 83.4 days (baseline) to 55.5 days. The scenario that is next best at reducing new cases is when 22% of agents self isolate after infectious period starts (65 day duration outbreak). The vaccination scenario (delivery to 50% of most high risk 30% of target group) produced negligible improvements over baseline, from 83.4 to 80.5 days and almost no reduction in new cases (17 to 16 median). The sensitivity analyses (Tables S1-S4) support the primary results.

**Table S1.** Sensitivity Analysis for Baseline scenario starting with 8, 16, 32 or 64 agents.

| **Simulation Outcomes** |  | **8 cases at start** | **16 cases at start** | **32 cases at start** | **64 cases at start** | **8, 16, 32, 64 agents** |
| --- | --- | --- | --- | --- | --- | --- |
| **Median: IQR** |  | **6 x 1000 simulations** | **6 x 1000 simulations** | **6 x 1000 simulations** | **6 x 1000 simulations** | **24,000 simulations** |
| Duration of outbreak from seeding to finish | days | 66.0: 39-141 | 97.3: 53-207 | 132.0: 67-267 | 170.0: 81-312 | 99.2: 52-234 |
|  | weeks | 9.4: 5.6-20.1 | 13.9: 7.6-29.7 | 18.9: 9.6-38.1 | 24.3: 11.5-44.6 | 14.1: 7-33 |
| Number of additional agents infected (secondary cases) by time outbreak finished |  | 8: 2-36 | 22: 6-103 | 56: 13-232 | 122: 28-476 | 28: 6-173 |

Note: Values are median : IQR. 6 transmission rate risks per encounter are: 6%, 12%, 24%, 36%, 48% or 60%. 1000 simulations were run for each of these 5 options in the baseline scenario conditions.

**Table S2.** Sensitivity analysis for partner reduction scenarios (1), starting with 8, 16, 32 or 64 agents.

| **Simulation Outcomes** |  | **8 cases at start** | **16 cases at start** | **32 cases at start** | **64 cases at start** |
| --- | --- | --- | --- | --- | --- |
| **Median: IQR** |  | **6 x 3 x 100 simulations** | **6 x 3 x 100 simulations** | **6 x 3 x 100 simulations** | **6 x 3 x 100 simulations** |
| Duration of outbreak from seeding to finish | days | 45.0: 34-72 | 58.0: 40-93 | 73.7: 49-118 | 89.1: 60-146 |
|  | weeks | 6.5: 4.9-10.3 | 8.3: 5.8-13.4 | 10.5: 6.9-16.8 | 12.6: 8.6-20.9 |
| Number of additional agents infected (secondary cases) by time outbreak finished |  | 4: 2-9 | 9: 4-22 | 19: 8-44 | 41: 17-96 |

Note: 5 values tested for risk of transmission per encounter are: 6%, 12%, 24%, 36%, 48% or 60%. Please see main text for description of partner reduction options (n=3). 100 simulations were run for each permutation and are pooled for this table.

**Table S3.** Sensitivity analysis for vaccination delivery rate scenarios (2), starting with 8, 16, 32 or 64 agents.

| **Simulation Outcomes** |  | **8 cases at start** | **16 cases at start** | **32 cases at start** | **64 cases at start** |
| --- | --- | --- | --- | --- | --- |
| **Median: IQR** |  | **6 x 3 x 100 simulations** | **6 x 3 x 100 simulations** | **6 x 3 x 100 simulations** | **6 x 3 x 100 simulations** |
| Duration of outbreak from seeding to finish | days | 63.5: 38-121 | 87.3: 51-157 | 116.0: 65-195 | 271.4: 234-323 |
|  | weeks | 9.1: 5.5-17.3 | 12.5: 7.4-22.4 | 16.4: 9.3-27.8 | 38.6: 33.5-46.1 |
| Number of additional agents infected (secondary cases) by time outbreak finished |  | 7: 2-29 | 19: 6-65 | 46: 13-144 | 504: 401-636 |

Note: Values are median: IQR. 6 transmission rate risks are: 6%, 12%, 24%, 36%, 48% or 60%. 3 vaccination roll out rates are: 25%, 50% or 75% of target population by day 120. 100 simulations were run for each permutation.

**Table S4.** Sensitivity analysis for self isolation scenarios (3), starting with 8, 16, 32 or 64 agents.

| **Simulation Outcomes** |  | **8 cases at start** | **16 cases at start** | **32 cases at start** | **64 cases at start** |
| --- | --- | --- | --- | --- | --- |
| **Median : IQR** |  | **6 x 4 x 100 simulations** | **6 x 4 x 100 simulations** | **6 x 4 x 100 simulations** | **6 x 4 x 100 simulations** |
| Duration of outbreak from seeding to finish | days | 48.0: 34-83 | 62.5: 40-115 | 86.3: 53-147 | 107.0: 63-184 |
|  | weeks | 6.9 : 4.9-11.9 | 9.9: 5.7-16.5 | 12.4: 7.6-21.1 | 15.4: 9.0-26.3 |
| Number of additional agents infected (secondary cases) by time outbreak finished |  | 4: 1-13 | 9: 3-33 | 26: 8-75 | 59: 15-159 |

Note: 6 transmission rate risks are: 6%, 12%, 24%, 36%, 48% or 60%. 4 proportions of population that self isolate after illness onset: 11%, 22%, 33% or 44%. 100 simulations were run for each permutation.

## Discussion

All of the strategies considered in our models (partner reduction, vaccination, self-isolation after illness onset) could be very effective to stop mpox outbreaks if deployed fully. However, reality is that behaviour change or vaccine uptake is often imperfect and takes time to implement. With respect to outbreak duration and new cases after simulation start, under plausible assumptions, our modelling suggests that reducing partner concurrency and casual partnerships were likely to have had the most benefits in reducing mpox spread (scenario 1). It may be surprising that smallpox vaccination had negligible effects on outbreak length and total number of secondary infections in our modelling (scenario 2). This finding likely arose because of the pace of vaccine delivery: only 50% administered to target group after four months (in reality and in the models). Faster vaccination delivery would have made the real-world vaccination strategy more effective. Free text comments about mpox collected as part of a 2022 survey of MSMs in the UK ^30^ indicated that many respondents were frustrated about or had encountered many difficulties in obtaining a smallpox vaccine.

Self-isolation (scenario 3) was much more beneficial than vaccination, but less effective than reductions in casual partnerships (scenario 1) in our models. Information about actual self-isolation adherence behaviour when infectious during the mpox epidemic is still emerging. Given the well documented challenges many people had in adhering to self-isolation requirements during the Covid19 pandemic ^32^, adherence to mpox self-isolation recommendations seems likely to have been low, perhaps much lower than our baseline choice of 22%. However, because close contact is generally required to transmit mpox, the self-isolation undertaken in reality may have been adequate if not ideal: people refrained from physical contact although they didn’t self-isolate in their homes entirely.

The World Health Organization alerted global authorities to the 2022 mpox outbreak in late May 2022 ^41^. The modelling here provides evidence for a common hypothesis ^42^ that the peak in case counts (estimated July 9 in UK ^43^ not long after this announcement and case count decline since, should be credited mostly to behaviour choices by members of the MSM community rather than vaccination programmes. Indeed, smallpox vaccination to MSMs at high risk of acquiring mpox in the UK was not widely offered before early July 2022 ^4^. Our modelling may be helpful in confirming the likely valuable contribution that personal behaviour choices have made and can continue to make in bringing mpox outbreaks under control. The importance of personal behaviour choices can be vital to share in public health campaigns. This statement is not intended to undermine the huge value of vaccination programmes; vaccines can confer lasting immunity and resistance to disease, and thus are a more reliable population protection measure than relying on individual behaviour choices. Protective sexual behavioural choices are often not sustained over long time periods ^44^. Ongoing smallpox vaccination for British MSMs was suggested as a key reason there was no large resurgence in new mpox cases in the UK in 2023 ^45^.

Sensitivity analysis (Tables S1-S4) which allowed for different numbers of infected agents at baseline (8, 16, 32 or 64) and different transmission risks (from 6-60% per contact), do not change the overall finding that reducing casual partnerships was relatively more effective as a epidemic control action than vaccination or self-isolation behaviour. However, compared to the baseline and plausible alternative scenarios, our larger simulation sets have wider interquartile ranges for the key epidemic harm outcomes: additional cases and duration of the outbreaks. Higher uncertainty about epidemic protection, especially over a longer time period, can be expected to associate more with behavioural strategies (partner reduction or some self-isolation) than with vaccination.

### Limitations and Strengths

It was not feasible to test our models against real world data. We are unaware of any real world situation that we could compare with, where only one of the strategies described here was applied to control mpox in MSMs in 2022. We also note that a mpox epidemic would not last perpetually in a closed system model such as this one; there is some possibility that depletion of the most active susceptible population also helped to hasten the decline of mpox cases in Britain in 2022.

Models are simplified versions of a complicated and complex world. Our models consider a relatively sexually active population; in reality, a large number of people have relatively low counts of sex events and partners in their lives (lower than our models suggest ^30^). Post-exposure smallpox vaccination is thought to offer some protection against developing infectious illness; our models don’t allow for this possibility. We designed our models to reflect plausible behaviour but lack information to confidently know how representative all of these many behaviour assumptions were of most MSMs in Britain or elsewhere. For instance, although we modelled vaccine deployment at a speed in accordance with published information about the actual UK programme, we did not consider if behaviour change was itself also slowly implemented. It is possible that change in behaviour in England was much slower than our models assumed, and as a result neither self-isolation nor partner reduction but rather other unmeasured factors were responsible for the decline in new English mpox cases after July 2022.

Our models attempt to examine each strategy in isolation and does not consider potential synergistic effects. Nor do we allow for varying implementation of any scenario. For instance, in reality, the vaccination programme likely had a slow start, the average pace of delivery at start of period was slower than by the end, because infrastructure takes a little while to create. The constant rate of vaccination delivery that our models applied will have slightly over-estimate rated of early dose delivery.

Our approach is novel in that we are not aware of other ABMs that address mpox spread among MSMs. ABMs can be relatively more flexible and naturalistic compared to other modelling approaches ^46^, explicitly incorporating heterogeneity of agent attributes and behaviour choices ^47^. ABMs are designed to reveal emergent phenomena ^46^ and thus may provide novel insights to epidemic dynamics. Our models inherently incorporated assortive mixing which is a well-recognised feature of actual social networks ^48,49^ possible contact rates and plausible behaviour choices. We used published information to construct all of the control scenarios, two of the scenarios emulated published data about partner count reductions and vaccination delivery. These ABM advantages come with a price in that they are computationally exhaustive ^47^ and with our software platform, meant that the agent population needed to be restricted (to 6400 agents) to generate many strategy and scenario outcomes in a reasonable timeframe.

In reality, MSMs in communities affected by mpox in 2022 doubtless undertook a blend of behaviour choices that helped to reduce transmission. We have not tried to estimate what the optimal mix of multiple strategies would be. Clearly, combined strategies are more effective than any strategy in isolation; deploying a variety of control strategies for any future mpox outbreaks is clearly wise in the real world.

## Data Availability

All data produced in the present study are available upon reasonable request to the authors

## Declarations

### Conflict of interest

The authors declare that we have no conflict of interest.

Approval to use the data to undertake the research Not required as this is a modelling study

## References

1. ECDC. Factsheet for health professionals on mpox (monkeypox). Jun 1 2023. https://www.ecdc.europa.eu/en/all-topics-z/monkeypox/factsheet-health-professionals (accessed Jan 5 2024).

2. Sklenovská N, Van Ranst M. Emergence of monkeypox as the most important orthopoxvirus Infection in humans. Frontiers in Public Health 2018; 6.

3. Poland GA, Kennedy RB, Tosh PK. Prevention of monkeypox with vaccines: a rapid review. The Lancet Infectious Diseases 2022.

4. Mahase E. Monkeypox: Single dose of smallpox vaccine offers 78% protection, UKHSA reports. British Medical Journal 2022; 379.

5. Reynolds MG, Damon IK. Outbreaks of human monkeypox after cessation of smallpox vaccination. Trends Microbiol 2012; 20(2): 80–7.

6. Thornhill JP, Barkati S, Walmsley S, et al. Monkeypox virus infection in humans across 16 countries—April– June 2022. New England Journal of Medicine 2022; 387(8): 679–91.

7. Patel A, Bilinska J, Tam JC, et al. Clinical features and novel presentations of human monkeypox in a central London centre during the 2022 outbreak: descriptive case series. Br Med J 2022; 378.

8. Sah R, Mohanty A, Abdelaal A, Reda A, Rodriguez-Morales AJ, Henao-Martinez AF. First Monkeypox deaths outside Africa: no room for complacency. Therapeutic Advances in Infectious Disease 2022; 9: 20499361221124027.

9. Vogel L. Making sense of monkeypox death rates. Can Med Assoc J 2022; 15(E1097).

10. Samarasekara K, Ringshall M, Parashar K, et al. Contribution of men who have sex with men (MSM) attending due to contact tracing to the diagnoses of HIV, syphilis and gonorrhoea in MSM from a clinic-based population. Sexually Transmitted Infections 2022; 98(4): 307–9.

11. Delaney KP. Strategies adopted by gay, bisexual, and other men who have sex with men to prevent monkeypox virus transmission—United States, August 2022. Morb Mortal Weekly Rep 2022; 71.

12. Cooney C, Durbin A. High-risk monkeypox contacts advised to isolate. BBC News. 2022 23 May.

13. Wellman MP. Putting the agent in agent-based modeling. Autonomous Agents and Multi-Agent Systems 2016; 30(6): 1175–89.

14. Pitt R, Foster C, Rayment M, et al. P132 Low prevalence of asymptomatic Mpox virus in men attending sexual health services in England. UK Health Security Agency Annual Meeting,; 2023; Leeds UK: BMJ Publishing Group Ltd; 2023.

15. Endo A, Murayama H, Abbott S, et al. Heavy-tailed sexual contact networks and monkeypox epidemiology in the global outbreak, 2022. Science 2022; 378(6615): 90–4.

16. Shalizi CR, Thomas AC. Homophily and contagion are generically confounded in observational social network studies. Sociological Methods & Research 2011; 40(2): 211–39.

17. Mac Carron P, Kaski K, Dunbar R. Calling Dunbar’s numbers. Social Networks 2016; 47: 151–5.

18. Brainard J, Hunter PR, Hall I. An agent-based model about the effects of fake news on a norovirus outbreak Rev Epidemiol Sante Publique 2020; 68(1): 99–107.

19. Mossong J, Hens N, Jit M, et al. Social contacts and mixing patterns relevant to the spread of infectious diseases. PLoS Med 2008; 5(3): e74.

20. Brainard J, Potts HW, Smith LE, Rubin GJ. The relationship between age and sex partner counts during the mpox outbreak in the UK, 2022. (under review) 2023.

21. Brainard J, Smith LE, Potts HW, Rubin GJ. The relationship between age and sex partner counts during the mpox outbreak in the UK, 2022. PLoS One 2023; 18(9): e0291001.

22. Pal M, Mengstie F, Kandi V. Epidemiology, diagnosis, and control of monkeypox disease: a comprehensive review. Am J Infect Dis Microbiol 2017; 5(2): 94–9.

23. Centers for Disease Control. Clinical Recognition: Key Characteristics for Identifying Mpox. 2023. https://www.cdc.gov/poxvirus/mpox/clinicians/clinical-recognition.html (accessed May 17 2023).

24. Adler H, Gould S, Hine P, et al. Clinical features and management of human monkeypox: a retrospective observational study in the UK. The Lancet Infectious Diseases 2022; 22(8): 1153–62.

25. Manzulli V, Scioscia G, Giganti G, et al. Real time PCR and culture-based virus isolation test in clinically recovered patients: is the subject still infectious for SARS-CoV2? Journal of Clinical Medicine 2021; 10(2): 309.

26. Ježek Z, Grab B, Szczeniowski M, Paluku K, Mutombo M. Human monkeypox: secondary attack rates. Bulletin of the World Health Organization 1988; 66(4): 465.

27. Besombes C, Mbrenga F, Malaka C, et al. Investigation of a mpox outbreak in Central African Republic, 2021-2022. One Health 2023; 16: 100523.

28. Nolen LD, Osadebe L, Katomba J, et al. Extended human-to-human transmission during a monkeypox outbreak in the Democratic Republic of the Congo. Emerging Infectious Diseases 2016; 22(6): 1014.

29. Office for National Statistics. Sexual orientation, UK: 2019. https://www.ons.gov.uk/peoplepopulationandcommunity/culturalidentity/sexuality/bulletins/sexualidentityuk/2019: Office for National Statistics, 2021.

30. Smith LE, Potts HW, Brainard JS, et al. Mpox knowledge, attitudes, beliefs, and intended behaviour in the general population and men who are gay, bisexual, and who have sex with men. medRxiv 2022.

31. Wang H, de Paulo KJdA, Gültzow T, Zimmermann HM, Jonas KJ. Monkeypox self-diagnosis abilities, determinants of vaccination and self-isolation intention after diagnosis among MSM, the Netherlands, July 2022. Eurosurveillance 2022; 27(33): 2200603.

32. Smith LE, Potts HW, Amlôt R, Fear NT, Michie S, Rubin GJ. Adherence to the test, trace, and isolate system in the UK: Results from 37 nationally representative surveys. British Medical Journal 2021; 372: n608.

33. Miura F, van Ewijk CE, Backer JA, et al. Estimated incubation period for monkeypox cases confirmed in the Netherlands, May 2022. Eurosurveillance 2022; 27(24): 2200448.

34. Guzzetta G, Mammone A, Ferraro F, et al. Early Estimates of Monkeypox Incubation Period, Generation Time, and Reproduction Number, Italy, May-June 2022. Emerging Infectious Diseases 2022; 28(10).

35. World Health Organization. Mpox (monkeypox). 18 Apr 2023. https://www.who.int/news-room/fact-sheets/detail/monkeypox (accessed 9 Sept 2023).

36. Prasad S, Casas CG, Strahan AG, et al. A dermatologic assessment of 101 mpox (monkeypox) cases from 13 countries during the 2022 outbreak: Skin lesion morphology, clinical course, and scarring. Journal of the American Academy of Dermatology 2023; 88(5): 1066–73.

37. Wellings K, Palmer MJ, Machiyama K, Slaymaker E. Changes in, and factors associated with, frequency of sex in Britain: evidence from three National Surveys of Sexual Attitudes and Lifestyles (Natsal). British Medical Journal 2019; 365.

38. Mercer CH, Jones KG, Geary RS, et al. Association of timing of sexual partnerships and perceptions of partners’ Concurrency with reporting of sexually transmitted infection diagnosis. JAMA Network Open 2018; 1(8): e185957-e.

39. Scott N, Stoové M, Wilson DP, et al. Eliminating hepatitis C virus as a public health threat among HIV-positive men who have sex with men: a multi-modelling approach to understand differences in sexual risk behaviour. J Int AIDS Soc 2018; 21(1): e25059.

40. Romero-Severson E, Volz E, Koopman J, Leitner T, Ionides E. Dynamic variation in sexual contact rates in a cohort of HIV-negative gay men. Am J Epidemiol 2015; 182(3): 255–62.

41. Organization WH. Multi-country monkeypox outbreak in non-endemic countries. May 21 2022. https://www.who.int/emergencies/disease-outbreak-news/item/2022-DON385 (accessed Dec 2 2022).

42. Kupferschmidt K. Monkeypox cases are plummeting. Scientists are debating why. 26 Oct 2022. https://www.science.org/content/article/monkeypox-cases-are-plummeting-scientists-are-debating-why (accessed Dec 4 2022).

43. Ward T, Christie R, Paton RS, Cumming F, Overton CE. Transmission dynamics of monkeypox in the United Kingdom: contact tracing study. Br Med J 2022; 379.

44. Sherman DW, Kirton CA. The experience of relapse to unsafe sexual behavior among HIV-positive, heterosexual, minority men. Appl Nurs Res 1999; 12(2): 91–100.

45. Zhang X-S, Mandal S, Mohammed H, et al. Transmission dynamics and effect of control measures on the 2022 outbreak of mpox among gay, bisexual, and other men who have sex with men in England: a mathematical modelling study. The Lancet Infectious Diseases 2023.

46. Bonabeau E. Agent-based modeling: Methods and techniques for simulating human systems. Proceedings of the National Academy of Sciences 2002; 99(suppl_3): 7280–7.

47. Molla J, Sekkak I, Ortiz AM, Moyles I, Nasri B. Mathematical modeling of mpox: A scoping review. One Health 2023: 100540.

48. Krzyżanowska M, Mascie-Taylor CN. Educational and social class assortative mating in fertile British couples. Ann Hum Biol 2014; 41(6): 561–7.

49. Phillips G, Birkett M, Hammond S, Mustanski B. Partner preference among men who have sex with men: potential contribution to spread of HIV within minority populations. LGBT health 2016; 3(3): 225–32.

